# GABAergic regulation of auditory repetition suppression in adults with and without Autism Spectrum Disorder

**DOI:** 10.1101/2023.02.15.23285928

**Authors:** Qiyun Huang, Hester Velthuis, Andreia C. Pereira, Jumana Ahmad, Samuel F. Cooke, Claire L. Ellis, Francesca M. Ponteduro, Nicolaas A. J. Puts, Mihail Dimitrov, Dafnis Batalle, Nichol M. L. Wong, Lukasz Kowalewski, Glynis Ivin, Eileen Daly, Declan G. M. Murphy, Gráinne M. McAlonan

**Author notes:** Corresponding author Corresponding author: Professor Gráinne M. McAlonan, Institute of Psychiatry, Psychology and Neuroscience (IoPPN), King’s College London, 16 De Crespigny Park, Denmark Hill, London, SE5 8AF. These authors contributed equally to this work (Declan G. M. Murphy, Gráinne M. McAlonan).

## Abstract

Suppressing responses to repetitive sounds, while staying vigilant to rare sounds, is a cross-species trait vital for survival, which is altered in autism spectrum disorder (ASD). Preclinical models implicate ϒ-aminobutyric acid (GABA) in this process. Although differences in GABA genes, post-mortem markers and bulk tissue GABA levels have been observed in ASD, the link between GABA and auditory processing in humans (with or without ASD) is largely correlational. Here, we directly evaluated the role of GABA in auditory repetition suppression in 66 adults (n = 28 with ASD). Neurophysiological responses (temporal and frequency domains) to repetitive standard tones and novel deviants presented in an oddball paradigm were compared after double-blind, randomized administration of placebo, 15 or 30 mg of arbaclofen (STX209), a GABA type B (GABA_B_) receptor agonist. We first established that temporal mismatch negativity was comparable between control participants and those with ASD. Next, we showed that temporal and spectral responses to repetitive standards were suppressed relative to responses to deviants in the two groups, but suppression was significantly weaker in individuals with ASD at baseline. Arbaclofen reversed weaker suppression of spectral responses in ASD but disrupted suppression in controls. An individual ‘sensitivity index’ of arbaclofen-elicited shift in suppression strongly correlated with autistic symptomatology measured using the Autism Quotient. Thus, our results confirm: GABAergic dysfunction is fundamental to the neurophysiology of auditory sensory processing alterations in ASD, which can be modulated by targeting GABA_B_ activity; and these GABA-dependent sensory differences may be upstream of more complex autistic phenotypes.

**One Sentence Summary:** Differences in GABAergic function are fundamental to autistic (auditory) sensory neurobiology; but are modulated by targeting GABA_B_.

## Introduction

Altered sensory reactivity, interests and/or responses in autism spectrum disorder (ASD) are now recognized as core to this diagnosis (Diagnostic and Statistical Manual of Mental Disorders, Fifth Edition) (*1*). Indeed, sensory features have been reported to be among the earliest indicators for ASD (*2, 3*). These features are prominent in the auditory domain and include both hyper- and hypo-responsivity to sounds: for example, excessive and adverse reactions to unexpected loudness (*4, 5*) and reduced orientation to ‘motherese’ (the profile of speech usually directed to, and preferred by, infants) (*6*). Such behavioral sensory features are assumed to be under-pinned by altered processing of sensory signals (*7-9*) and preclinical studies have implicated alterations in the inhibitory γ -aminobutyric acid (GABA) system (*10-12*). However, until recently (*13*), human studies have largely relied on correlational evidence for relationships between GABA genes, post mortem markers and bulk tissue GABA concentrations with sensory functions in ASD (*14, 15*). There is no direct experimental evidence that modulating the GABA system differentially modulates the processing of auditory information in the living human brain of individuals with and without ASD.

The brain continually adapts to sensory inputs to filter out irrelevant stimuli and aid detection and response to deviant stimuli that may be meaningful, for example, those signaling danger or reward (*16*). In the auditory domain, neurophysiological responses to sounds adapt (reduce) to repeated, predictable sounds but increase to novel, unpredictable stimuli (*17, 18*). This ‘mismatch’ is fundamental for auditory perception as repeated inputs are sifted out or deprioritized, whereas responses to novel signals are facilitated. A widely used cross-species model for studying this mismatch between repeated and novel stimuli in the auditory domain is the oddball paradigm, in which a train of identical repeated sounds (standards) is randomly interrupted by an oddball sound (deviant). Electroencephalogram (EEG) recording then captures responses to standards and deviants and the difference between standard-deviant response is termed mismatch negativity (MMN) (*19-23*).

Though the specific molecular or circuit mechanisms supporting MMN remain uncertain, there is consensus that at least two mechanisms are essential for its generation:

1. Stimulus-specific adaptation (SSA) in animals or repetition suppression in humans, which allows neural responses to adapt (reduce) to repeated sounds while maintaining responsiveness to deviants (*24*).
2. Deviance detection, which allows generation of increased responses to novel stimuli.

MMN generation has been explained under a predictive coding framework of Bayesian perceptual inference (*25, 26*) in which the auditory system is hierarchically organized. Each level sends ‘back’ predictions to aid the suppression of ascending neuronal activity evoked by anticipated sounds at downstream levels and sends ‘forward’ a prediction error signal to upstream levels when a failure to predict bottom-up information happens. An alternative explanation is that selective feedforward adaptive filtration occurs for synapses that mediate the familiar stimulus but not for those synapses that process the novel stimulus (*27, 28*). Under either framework, repetition suppression and deviance detection must co-exist; and this has been verified by decomposing the auditory mismatch response in preclinical animals(*29*).

In humans, neurodevelopmental conditions such as ASD alter brain function across the subcortical and cortical systems known to be involved in sensory processing from birth(*30, 31*). However, results from the oddball paradigm in ASD are heterogeneous. A majority of studies have limited analysis of the oddball paradigm to the event-related response MMN. Smaller amplitudes and/or delayed latencies (*32, 33*), or no differences (*34*) in MMN have been reported in individuals with ASD relative to individuals with typical development (TD). A meta-analysis proposed that age partially accounts for this variability, such that children with ASD tend to have smaller MMN amplitude relative to TD, whereas adult groups have comparable measures (*35*).

However, inconsistent results from the oddball paradigm in ASD could also be a consequence of the multi-source distribution of MMN generators and the metric used. Both repetition suppression and deviance detection have been observed in neurons along the hierarchical auditory pathway, including cortical regions, such as the auditory cortex and medial prefrontal cortex, and subcortical nuclei like the inferior colliculus and the medial geniculate body (*36-38*). Work in rodents suggests that these two components exert varying degrees of influence on the MMN depending on the specific brain region (*29*). Repetition suppression is most prominent in the subcortical components and can be pharmacologically modulated in preclinical animals by targeting GABA signaling pathways (*39, 40*). In contrast, deviance detection is most prominent at high level cortical regions (*29*) and is modulated by N-methyl-D-aspartate receptor activity (*41-43*). Although scalp EEG is the resultant of multiple sub-cortical and cortical sources, the MMN, as quantified by subtracting the standard event-related potential (ERP) from the deviant ERP, is thought to best reflect cortical-based deviance detection. Moreover, this conventional event-related temporal MMN amplitude measurement provides only a brief snap-shot of the response to auditory stimulation. Therefore, perhaps unsurprisingly, results from studies focusing solely on event related MMN amplitudes are inconsistent as they have limited capture of the components operating during processing of repeated and novel auditory stimuli.

In contrast, time-frequency analysis, namely Event Related Spectral Perturbation (ERSP) (*44*), assesses changes in under-lying neuro-oscillatory dynamics in response to standards and deviants without subtraction (*45-47*). Neuro-oscillatory activity provides a foundation for the functional connectivity across brain regions which supports cognitive and behavioral output (*48*). Neuronal oscillations in specific frequency bands have been linked to GABAergic cellular characteristics (*49, 50*). These observations may be of critical relevance for ASD given that altered functional connectivity across brain networks is a replicable feature of the condition (*51*) and GABAergic differences are frequently reported in this condition (*52*). However, no-one has directly tested the hypothesis that alterations in the neuro-oscillatory responses to auditory stimuli are underpinned by dysfunctions in the GABAergic system in ASD; therefore this was an important goal of our work.

Here we investigated how altering GABA function modulates auditory processing in individuals with and without ASD. Scalp EEG was used to record neurophysiological responses induced by a classical auditory oddball paradigm in which a stream of regular repeating standard sounds was interrupted by oddball sounds that deviate from standards in frequency, duration or frequency-duration combination (*53*). Participants were tested at placebo or after a single oral dose of 15 mg or 30 mg arbaclofen (STX209), a selective GABA type B (GABA_B_) receptor agonist that has been demonstrated to be safe and well tolerated in ASD (*54, 55*). First, a conventional event-related potential (ERP) analysis was implemented to compare the MMN between TD and ASD at different drug administrations. Second, to obtain a more comprehensive capture of auditory processing in the oddball paradigm, an ERSP time-frequency analysis of responses to standard tones and deviants without subtraction, was a key focus of our analyses.

Given the evidence for higher neural reactivity (weaker suppression) to repetitive stimuli in children with ASD and infants at-risk of ASD (*2, 56, 57*) and reports of ASD-related alterations in the GABA system (*10, 13, 15*), we had two main predictions: (i) Repetition suppression reflected by oscillatory responses to standard tones would be atypical in ASD relative to TD; (ii) Repetition suppression would be differentially modulated by challenging the GABA system in ASD and TD. In contrast, we did not expect to find significant case-control differences in the event-related MMN signal in this adult cohort, nor any statistically significant modulation of ERPs in response to GABA_B_ agonism.

## Results

### Clinical cohort

Thirty-eight TD (22 males, 16 females) and 28 ASD (20 males, 8 females) were included and 138 study visits were completed: 50 placebo (P) visits (26 TD, 24 ASD), 50 low-dose (L) visits (30 TD, 20 ASD), and 38 high-dose (H) visits (19 TD, 19 ASD). Please see Table 1 for details of participant demographic characteristics.

**Table 1.** Participant demographic data and ASD clinical scores

| Measure | TD | ASD | Statistic | p |
| --- | --- | --- | --- | --- |
| Number<br>(male / female) | 22 / 16 | 20 / 8 | $\chi^2 = 1.3$ | 0.26 |
| Age | 28.6 ± 8.1 | 34.8 ± 10.1 | t = 2.7 | 0.01 |
| Full-scale IQ | 120.2 ± 10.5<sup>a</sup> | 117.4 ± 10.4<sup>b</sup> | t = 1 | 0.32 |
| AQ | 16.9 ± 8.1<sup>c</sup> | 35.1 ± 7.5<sup>d</sup> | t = 9.1 | 7 × 10<sup>-13</sup> |

Values are shown as means ± SD. Statistical results for the group difference of age, IQ and AQ (Autism Quotient) scores are the outcome of independent-sample t tests; and comparison of proportion of males and females carried out using Chi-squared test. As a result of Covid lockdown restrictions and personal unwillingness it was not possible to complete in-person IQ testing on <sup>a</sup>two neurotypicals; <sup>b</sup>six autistics. AQ data not returned for <sup>c</sup>three neurotypicals; <sup>d</sup>one autistic.

### MMN was comparable in ASD and TD; arbaclofen had a minimal impact on the MMN in the TD group only

The MMN is a difference waveform calculated by subtracting the averaged response to standard stimuli from averaged responses to each of the three deviants: frequency deviant, duration deviant and combined frequency-duration deviant (see Materials and Methods). As shown in Fig. 1A, the MMN appeared as a negative valley in the grand average waveforms within the interval [100, 250] ms post stimulus onset and persisted across stimulus conditions and drug administrations in both groups. Individual-level ERP features including the MMN amplitudes and the corresponding latencies are shown in Fig. 1B.

**Fig. 1.**
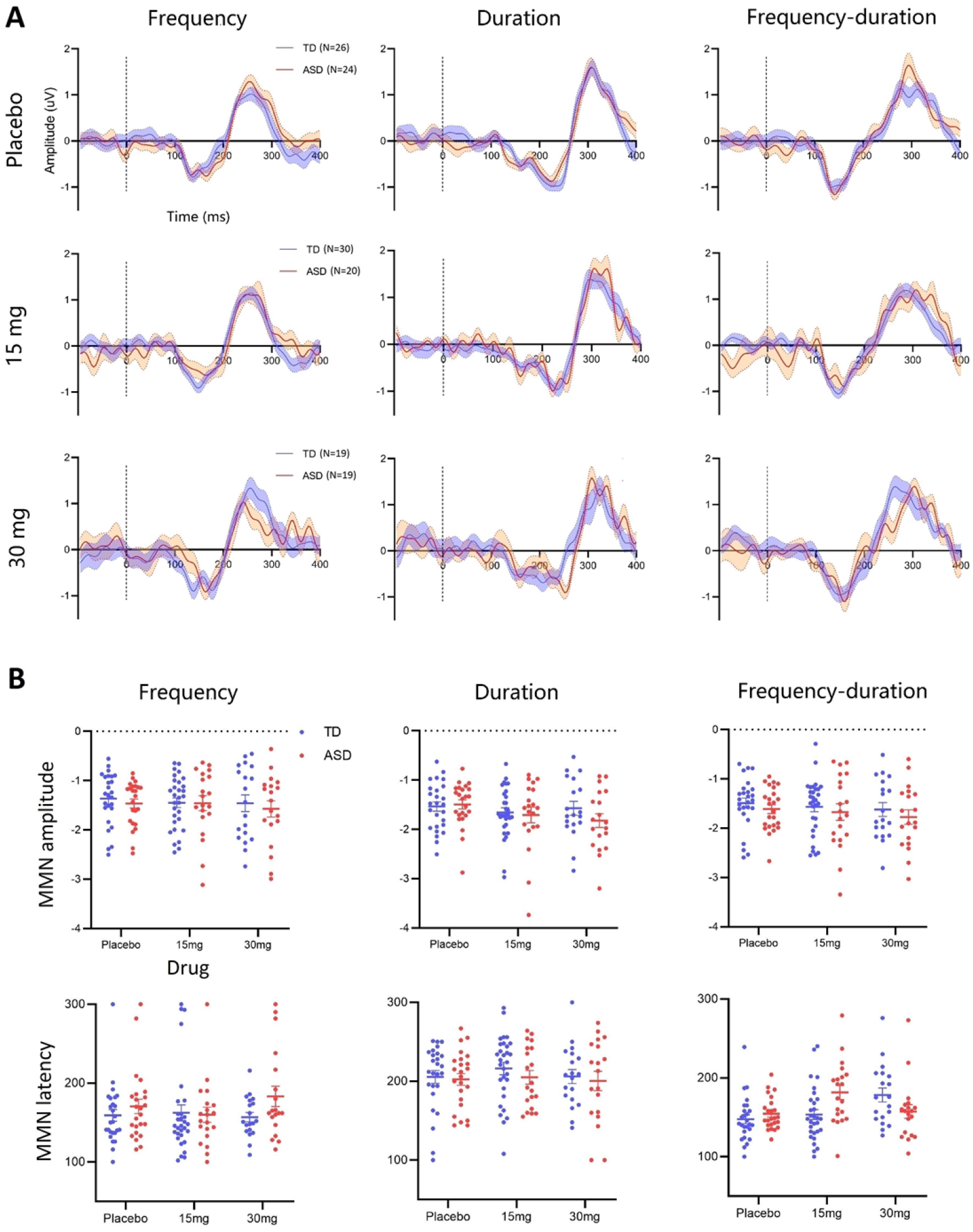
ERP waveforms and individual features of MMN. (A) The MMN grand average waveforms of the TD group (blue) and ASD group (red) as functions of the stimulus conditions (column) and drug administrations (row). Data epoch was drawn in the interval [-100, 400] ms referenced to the stimulus onset at 0 ms. Dashed lines indicate the stimulus onsets. (B) Individual scatter plots of MMN amplitudes (uV) and latencies (ms). *N*, number of participants. Error bar shows the standard error of the mean (SEM).

We used linear mix-effect models (LMM) to perform a repeated measures analysis examining the effect of deviant characteristic, group difference and drug effect on the individual MMN amplitudes and latencies. The p-values reported were corrected for multiple comparisons using Benjamini-Hochberg method (*58*). Please see Materials and Methods for details of statistical analyses.

*MMN amplitud*es: Across the whole cohort, significant differences were observed between frequency-MMN and duration-MMN (t_(409)_ = -3.1, p = 0.004), frequency-MMN and combined-MMN (t_(409)_ = -2.8, p = 0.006), but not between duration-MMN and combined-MMN (t_(409)_ = 0.2, p = 0.8). There were no significant effects of group or drug nor any interactions in the MMN amplitudes in any of the three deviant stimulus conditions.

*MMN latencie*s: Across the whole cohort, significant differences were observed between duration-MMN and frequency-MMN (t_(409)_ = -8.5, p = 3.3×10^−16^), duration-MMN and combined-MMN (t_(409)_ = -9.4, p = 3.8×10^−19^), but not between frequency-MMN and combined-MMN (t_(409)_ = -0.9, p = 0.5). This pattern was expected as the duration deviant must necessarily alter MMN latencies. A significant drug effect on the latencies of the combined-MMN was found in TD (t_(73)_ = 3, p = 0.02) but not in ASD (t_(73)_ = 0.4, p = 0.7); but neither the group effect (difference) nor the group-drug interaction reached statistical significance. No group difference, drug effect or interaction was found for the frequency-MMN or duration-MMN.

### Repetition suppression in ERP responses occurred in both TD and ASD; but suppression was significantly less in ASD

Standalone grand average ERP waveforms of responses to the standard tones and the three deviants (frequency, duration and frequency-duration) without subtraction are shown in Fig. 2A. Three prominent ERP components were observed in both the TD group and the ASD group: (i) an early P1 component appeared as a positive peak within the range [50, 100] ms post stimulus onset; (ii) a negative N1 components located within [100, 200] ms; (iii) a late P2 component followed N1 as a positive peak. The P2 component mostly contributed to the P3a after the MMN (Fig. 1A) and was therefore not further examined. For the duration deviant, there were two negative valleys located within the range [100, 300] ms and the latter was used as it was more aligned with the temporal range of the duration MMN (Fig. 1A). Please see Materials and Methods for details of the ERP feature extractions.

**Fig. 2.**
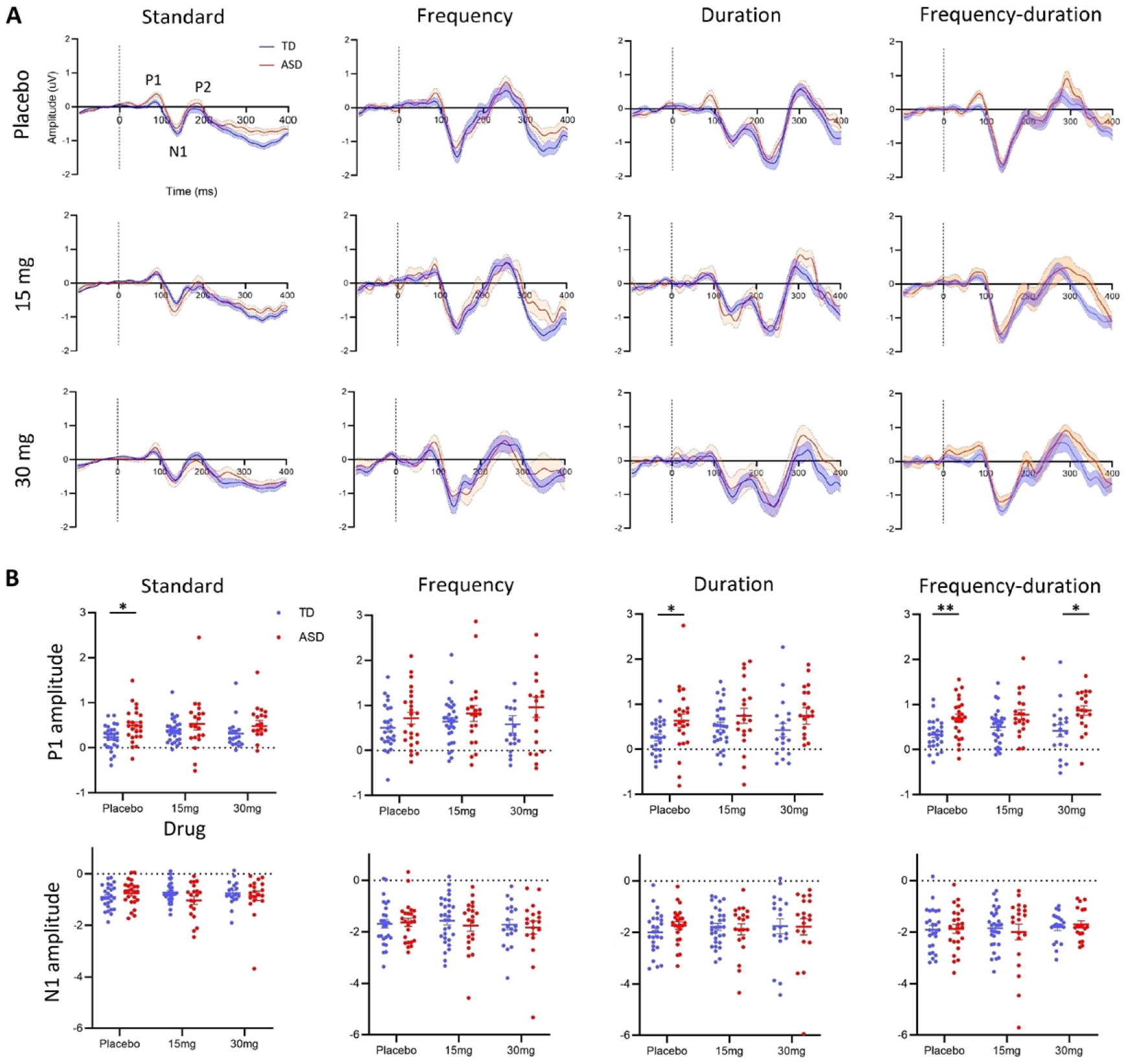
ERP waveforms and individual features of responses to standard tones and deviants. (A) The grand average waveforms of ERP responses to standard tones and the three deviants (frequency, duration and combined) without subtraction at the three drug administrations (row). Dashed lines indicate the stimulus onsets. (B) Individual scatter plots of the P1 and N1 amplitudes (uV) for the TD (blue) and ASD (red) group as a function of drug administration at the four stimulus conditions. *, the ASD-TD group difference is significant; corrected p < 0.05. **, the ASD-TD group difference is significant; corrected p < 0.01.

Individual scatter plots of the amplitudes of the P1 component are shown in Fig. 2B (top). The P1 amplitudes of repeated standard tones were significantly attenuated (suppressed) relative to any of the three deviants in the whole cohort: frequency (t_(546)_ = 5.5, p = 5.8×10^−8^), duration (t_(546)_ = 2.7, p = 0.01), and combined deviant (t_(546)_ = 3.4, p = 0.002). There was also a significant difference between response to frequency and duration deviants (t_(546)_ = -2.8, p = 0.01) and the difference between response to frequency and combined deviants was on the threshold of significance (t_(546)_ = -2, p = 0.05). There was no difference between duration and combined deviants (t_(546)_ = 0.7, p = 0.4). In the placebo condition, the ASD group showed significantly higher P1 amplitudes than TD in response to standard tones (t_(48)_ = 2.8, p = 0.01), duration deviants (t_(48)_ = 2.3, p = 0.03) and combined deviants (t_(48)_ = 3.3, p = 0.004); the mean of ASD in response to frequency deviants was also higher than TD though this difference was not significant (t_(48)_ = 1.3, p = 0.2). No drug effect was observed for TD or ASD at any conditions.

***Thus, the autistic brain was significantly ‘over-responsive’ to standard and deviant stimuli***.

Individual scatter plots of the amplitudes of the N1 component are shown in Fig. 2B (bottom). Similar to the P1, the N1 amplitudes of the responses to standard tones were significantly suppressed relative to any of the three deviants in the entire cohort: frequency (t_(546)_ = 11.1, p = 7.5×10^−26^), duration (t_(546)_ = 12.8, p = 3.2×10^−33^), and combined deviant (t_(546)_ = 13.3, p = 3.8×10^−35^). There was no difference in response between any pair of the three deviants. No group difference, drug effect or interaction was observed in any stimulus conditions.

### Repetition suppression of spectral responses was weaker in ASD; arbaclofen shifted spectral responses to a more typical profile in ASD but disrupted spectral response in TD

Brain dynamics of spectral responses to standard tones and the three deviants were captured using measures of event-related spectral perturbation (ERSP). ERSP was calculated in a single-trial manner and was used to measure dynamic changes induced by the stimuli in the broad band frequency spectrum as a function of time (see Materials and Methods).

#### Qualitative observations

The ERSP outcomes for the TD and ASD group under different stimulus conditions and drug administrations are presented in Fig. 3A and B, respectively. As revealed by the grand average in the placebo condition, repeated standard tones induced perturbations in the theta and alpha band [4, 12] Hz in ASD but spectral responses to repeated standards were clearly suppressed in TD. Low-dose (15 mg) and high-dose (30 mg) arbaclofen caused suppression of the standard-induced changes in ASD but reduced suppression in TD. The three deviants induced prominent spectral perturbations in both TD and ASD regardless of drug administration.

**Fig. 3.**
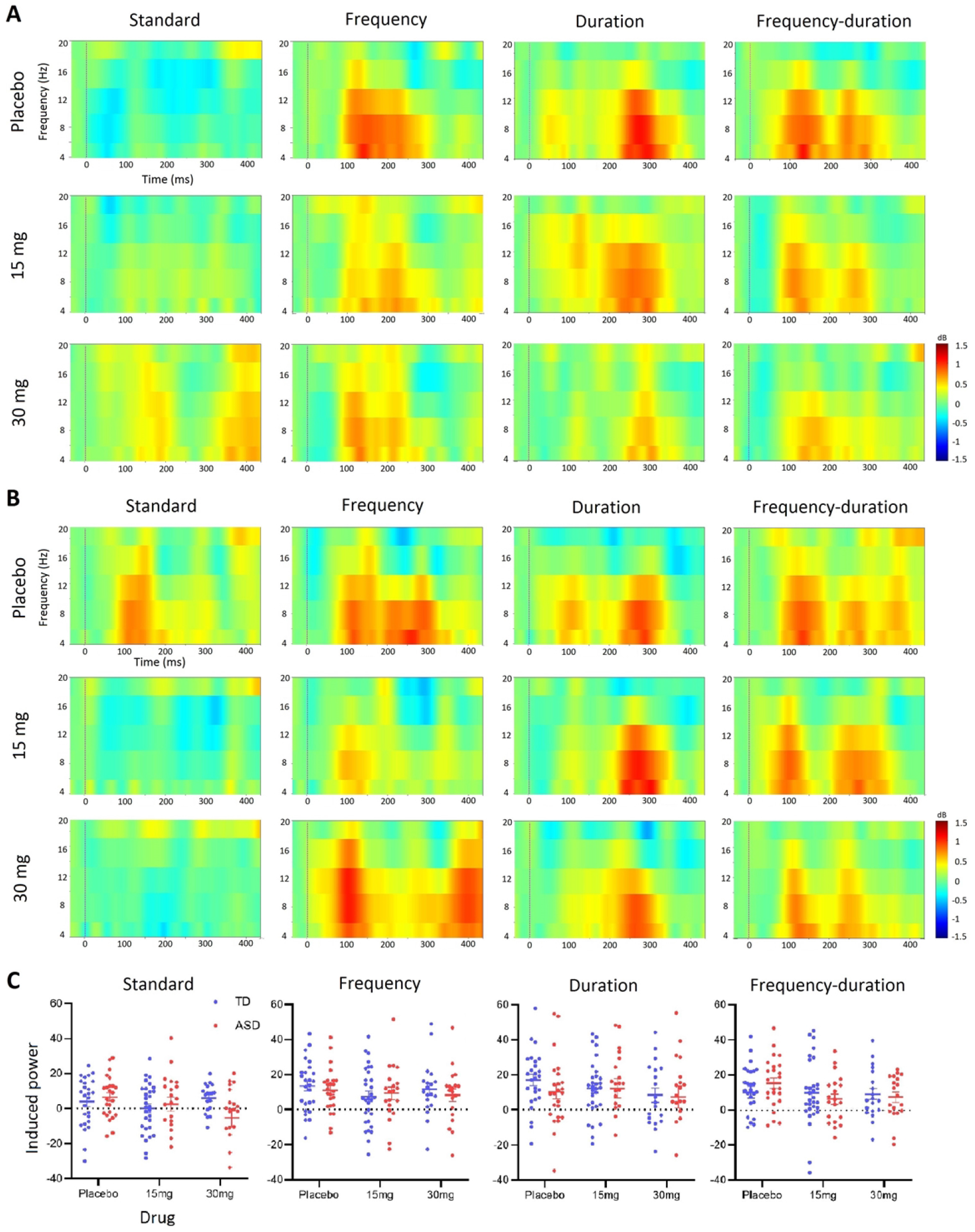
ERSP responses to standard tones and deviants. The grand average ERSP outcomes under different stimulus and drug conditions for the TD group (A) and the ASD group (B). Dashed lines indicate the stimulus onsets. (C) The scatter plots of individual measurements of spectral changes in [4, 12] Hz to standard tones and deviants as a function of drug administrations in TD (blue) and ASD (red) groups.

#### Quantitative observations

Scatter plots of the ERSP responses in the [4, 12] Hz band were extracted for statistical analysis and are shown in Fig. 3C. The spectral responses to repeated standard tones were significantly suppressed relative to any of the three deviants in the whole cohort: frequency (t_(546)_ = 3.9, p = 3×10^−4^), duration (t_(546)_ = 4.7, p = 3.5×10^−6^), and combined deviant (t_(546)_ = 3.7, p = 4×10^−4^). There was no difference between any pair of the three deviants. Although the group difference in spectral response to standards did not reach significance (t_(48)_ = 1.4, p = 0.1), there was a significant group-drug interaction (t_(134)_ = -2.1, p = 0.03) in responses to standard tones. This was explained by a significant drug effect in spectral response to standards in ASD (t_(61)_ = -2.3, p = 0.02) but not in TD (t_(73)_ = 0.4, p = 0.7). No group difference, drug effect or interaction was observed in responses to deviants.

***Thus, GABA***_***B***_ ***agonism reversed weaker suppression of spectral responses to repeated standards in ASD but did the opposite in TD***

However, this result may have been influenced by the number of times a standard is repeated between deviants; which is variable. Hence, to better understand repetition suppression post-hoc, we next examined the ERP and the spectral response to the standard just before and just after a deviant as these would typically be expected to be the most and least suppressed responses respectively.

### Post-hoc analyses

1. **Repetition suppression to pre-deviant standards was significantly weaker in ASD and was rescued by arbaclofen**.
2. **Repetition suppression to pre-deviant standards was stronger in TD and disrupted by arbaclofen**.

In a repeated sequence, brain responses to later stimuli are typically expected to be attenuated or suppressed by the repetition, while leaving responses to stimuli at the beginning of a sequence (after a deviant) less affected (*2*). We defined the ‘pre-deviant standard’ as the last sound in a four-in-a-row standard sequence before a deviant and the ‘post-deviant standard’ to be the first standard after a deviant (Fig. 4A). We examined both ERP and ERSP responses to the two types of standards for completeness (see Materials and Methods).

**Fig. 4.**
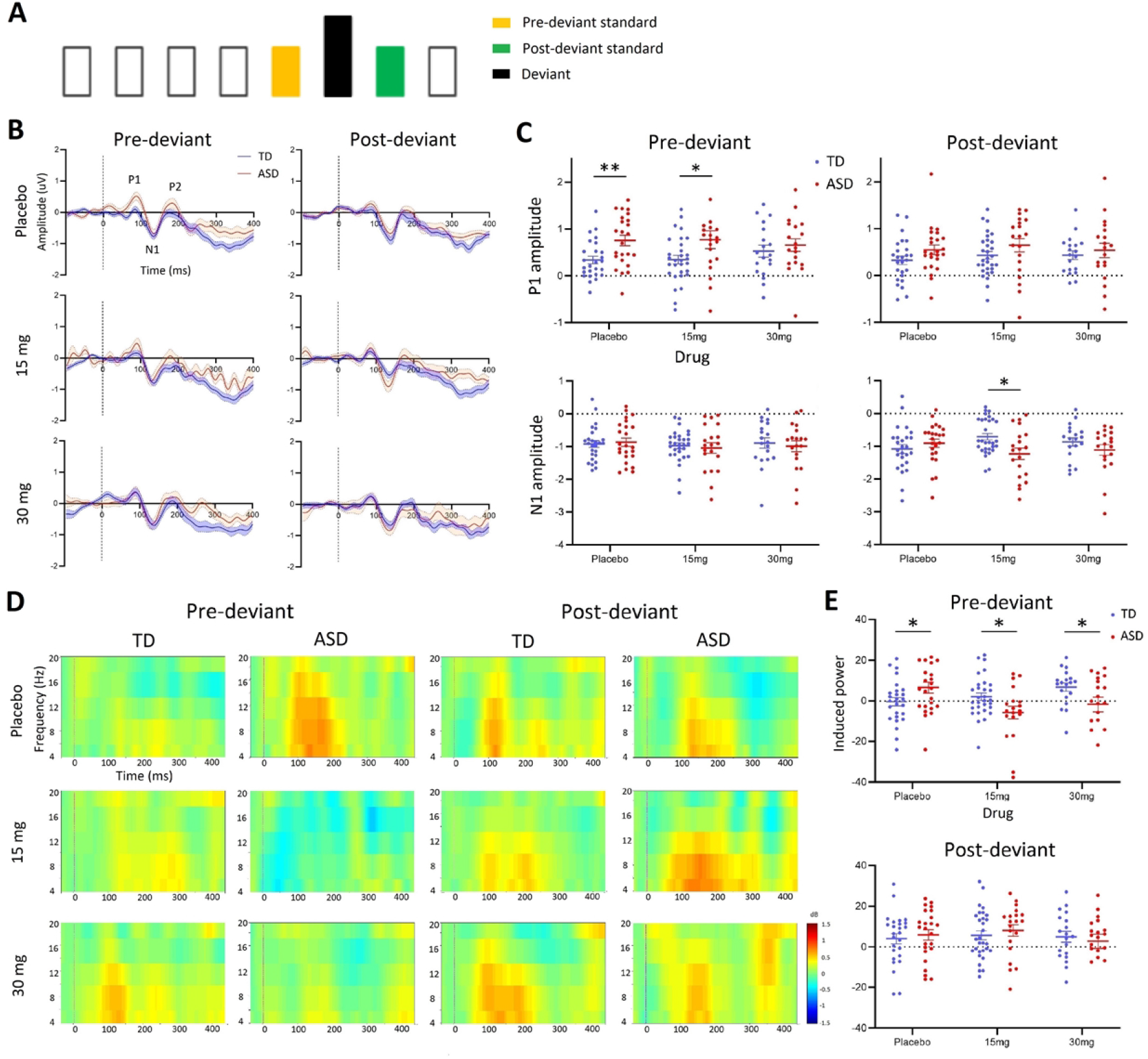
ERP and ERSP responses to pre-deviant and post-deviant standards. (A) Timeline schematic of an example stimulus delivery that the pre-deviant standard (yellow) and post-deviant standard (green) are adjacent to the deviant (black). (B) The grand average waveforms of ERP responses to pre-deviant and post-deviant standards at different drug administrations for TD (blue) and ASD (red). Dashed lines indicate the stimulus onsets. (C) Scatter plots of individual measures of ERP components. (D) The grand average ERSP responses to pre-deviant and post-deviant standards. (E) Scatter plots of individual measures of induced spectral responses. *, the ASD-TD group difference is significant; corrected p < 0.05. **, the ASD-TD group difference is significant; corrected p < 0.01.

#### ERP

The ERP waveforms and individual scatter plots of ERP components (P1 and N1) are shown in Fig. 4B and C, respectively. For P1 amplitudes, at placebo, individuals in the ASD group had significantly higher responses to pre-deviant standards relative to TD (t_(48)_ = 3.1, p = 0.009); at 15 mg, the ASD-TD group difference remained significant but with a less marked effect (t_(48)_ = 2.2, p = 0.04); at 30 mg, there was no difference between the two groups (t_(36)_ = 0.7, p = 0.5). These results supported modulation of P1 amplitudes by GABA_B_ receptor activation with arbaclofen, though LMM measures of drug effect or group-drug interaction did not reach significance. In contrast, there were no effects of group or drug on P1 amplitudes of the post-deviant standards. For N1 amplitudes, there were no significant differences between TD and ASD in neither the pre-deviant nor post-deviant conditions.

#### ERSP

The grand average ERSP responses to pre- and post-deviant standards are shown in Fig. 4D. This indicated that the ASD-TD difference in standard responses previously observed in the placebo condition (Fig. 3) was driven by differences in the pre-deviant but not the post-deviant responses. At individual level (Fig. 4E), significant ASD-TD differences were observed at all drug conditions in responses to pre-deviant standards (at placebo, t_(48)_ = 2.1, p = 0.04; at 15 mg, t_(48)_ = -2.3, p = 0.04; at 30 mg, t_(48)_ = -2.1, p = 0.04) but not to post-deviant standards. LMM results also confirmed that both drug (t_(133)_ = 3, p = 0.006) and group (t_(133)_ = 2.5, p = 0.02) had a significant effect on responses to pre-deviant standards but not to post-deviant standards. Moreover, a strong group-drug interaction was obtained for pre-deviant standards (t_(133)_ = -3.3, p = 0.002). Specifically, spectral responses to pre-deviant standards increased with drug dose in TD (t_(73)_ = 2.4, p = 0.01; weaker suppression with increasing dose) while they decreased in ASD (t_(60)_ = -2.3, p = 0.02; stronger suppression with increasing dose).

***Thus, GABA***_***B***_ ***activation with arbaclofen enhanced repetition suppression in ASD and disrupted it in TD through an action on standards prior to the deviant***.

### Individual sensitivity to GABA_B_ activation and relationship with autistic symptomatology

Finally, as ASD is a heterogeneous condition, we wished to understand GABA_B_ responses at the level of the individual as this may guide stratification in future clinical trials targeting GABA_B_ pathways. Therefore, we tested the hypothesis that an individual’s repetition suppression response to GABA_B_ receptor agonism would be correlated with the extent of their autistic features captured using the Autism Quotient (AQ; (*59*)).

Specifically, because the differential response to arbaclofen in TD and ASD groups was especially prominent in the spectral responses to pre-deviant standards, we defined a GABA_B_ ‘sensitivity index’ for each individual as the difference in spectral responses to pre-deviant standards at placebo minus those at 30 mg arbaclofen.

The placebo-30 mg transitions in the two groups are shown in Fig. 5A. Eleven out of 12 (92%) TD showed an increased effect of drug and resulted in a significant within-group placebo-30 mg difference (t_(11)_ = 5.1, p = 3.4×10^−4^). In contrast, 11 out of ASD (65%) showed a decreased effect of drug, still generating a significant within-group placebo-30 mg difference (t_(16)_ = -2.8, p = 0.01). The ASD-TD group difference in the sensitivity index was also significant (t_(27)_ = 4.4, p = 1.3×10^−4^; Fig. 5B). As predicted, there was a strong correlation between GABA_B_ response sensitivity and total scores on the AQ (r_(26)_ = -0.7, p < 0.0001) as shown in Fig. 5C; and between response to AQ question 5: “I often notice small sounds when others do not” (r_(26)_ = -0.6, p < 0.001) as shown in Fig. 5D. This indicated that the more atypical an individual’s GABA_B_-dependent auditory dynamics were, the more autistic features (total AQ scores) they had and the greater their perceptual sensitivity to sound was.

**Fig. 5.**
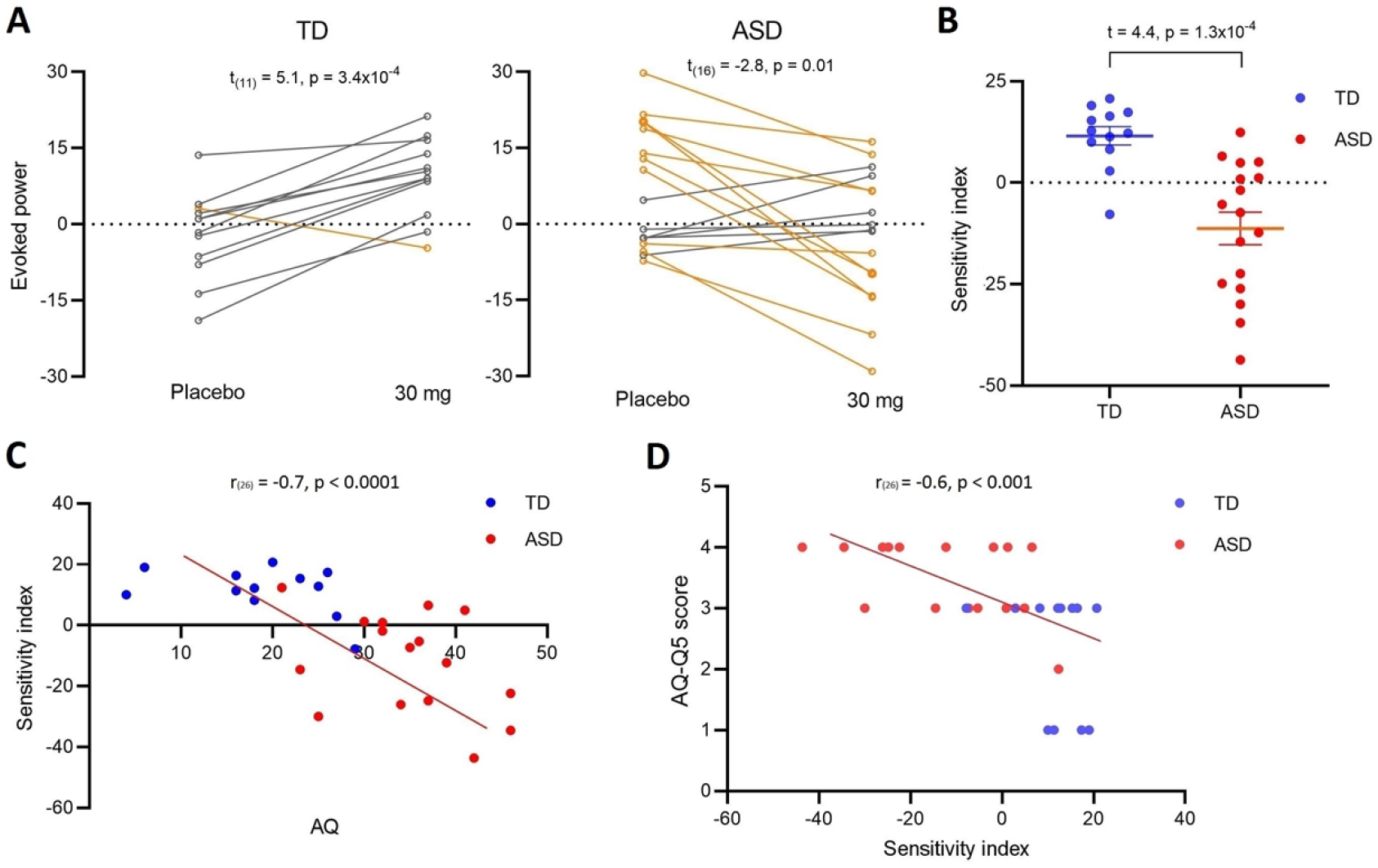
Individual sensitivity index and relationship with symptomatology. (A) The transition graphs to show the shift effect between placebo and 30 mg arbaclofen on TD and ASD. Yellow lines indicate a decrease in evoked power following arbaclofen (the majority of the ASD group); grey lines indicate an increase in evoked power following arbaclofen (the majority of the TD group). Only one individual in the TD group (yellow) had a decrease in evoked power in response to arbaclofen (behaving more like the ASD group); that person also had the highest AQ score in the TD group. (B) The scatter plots of extracted sensitivity indexes. (C) The correlation between the sensitivity index and total AQ scores. The point at which the direction of the sensitivity index changed (i.e. crosses the x-axis) approximates the AQ ‘cut-off’ for ASD (*59*). (D) The correlation between the sensitivity index and sound sensitivity captured by AQ question 5 score.

## Discussion

Differences in sensory processing are core to ASD, and these differences have been postulated to arise from alterations in excitation – inhibition balance (*60*), and especially GABAergic dysfunction (*61*). However, until now, no-one had directly tested the hypothesis in humans, that differential neural responses to auditory stimuli in ASD are underpinned by GABA.

In this study, we used an auditory odd-ball paradigm to confirm our two main predictions: (i) Time-frequency analyses revealed atypical repetition suppression in ASD relative to TD; (ii) Repetition suppression was differentially modulated by challenging the GABA system in ASD and TD. Specifically, we report that there is weaker suppression in both temporal ERP and spectral ERSP responses to repetitive standard tones in ASD relative to TD at placebo (baseline), which is consistent with prior ERP work (*2*). We then focused on the pre-deviant standards which were expected to be most affected by repetition suppression and demonstrated, for the first time, that activating GABA_B_ receptors through a single oral dose of arbaclofen could reverse atypical auditory processing in ASD and disrupt typical responses in TD. Thus, we have directly confirmed that that GABAergic dysfunction contributes to the neurophysiology of auditory alterations in ASD.

Importantly, we have also moved beyond group level approaches to examine the heterogeneity within the ASD and TD groups. We showed that the extent to which an individual responded to modulation of the GABA system with arbaclofen could be captured by their sensitivity index and this index strongly correlated with that person’s ASD symptomology. Moreover, when we extracted the AQ question capturing subjective sensitivity to sounds (perception), we found this also strongly correlated with GABAergic response. Thus, more autistic characteristics, including auditory perceptual features, as measured using the AQ were associated with greater recovery of repetition suppression in response to arbaclofen. Indeed, individuals with AQ scores above the 26 point ‘cut-off’ for autism spectrum (the majority of the ASD group) had weaker repetition suppression which increased following arbaclofen; individuals with AQ scores below 26 (the majority of controls) had stronger repetition suppression which was weakened by arbaclofen. Thus, GABA_B_ receptor activation has diametrically opposite effects on auditory processing in people who score above and below a phenotypic ‘cut-off’ for ASD. We cautiously suggest that this implies a normal distribution for auditory processing in the population. That is, individuals with ASD distant from the peak of this measure, but can be shifted towards peak processing by arbaclofen, while those close to peak auditory processing (majority of TD) at baseline are shifted away by arbaclofen. Further work will be required to confirm this hypothesis. Importantly, however, our work indicates that auditory processing profiles are not ‘fixed’ in either ASD or TD; they can be modulated, even in adults.

### Adapting to repeated and novel stimuli

Our results indicate that in ASD there is a relative failure to dampen the response to repetitive and predictable information (weaker repetition suppression), while preserving the response to true deviants (normal deviance detection). These results can sit comfortably within a predictive coding (*62*) or feedforward adaptive filtration framework (*27*), as repetition suppression is manifest in either case.

#### Event-related MMN

There was no group difference at baseline and minimal effect of arbaclofen in either group on the ERP-measured MMN. This is consistent with previous negative results in oddball studies of ASD because the ERP which is used to calculate the MMN is largely determined by deviance detection rather than response suppression (*34, 35*). This again can be explained within either predictive coding or feedforward adaptive filtering frameworks. The former theory postulates that the deviance detection captured by the MMN relies on the formulation of a short-term trace of previous regularity and is memory-based; it relies on the NMDA receptor, and may not need to be GABA-dependent (*63, 64*). NMDAR function is thought to be either intact or, at most, heterogeneous in ASD (*65*). Therefore, we might expect minimal group differences and/or response to GABAergic challenge on this metric. The latter theory of adaptive filtration only requires that synapses that are modified in response to the repeated stimulus are distinct from those that process the novel stimulus, so it allows for event-related MMN to continue in both groups and be relatively unchanged by arbaclofen.

#### Repetition suppression

Repetition suppression (SSA in preclinical animals), however, has been clearly demonstrated in preclinical studies to be modulated by the GABA system (*39, 40*). Thus, our observations indicate that findings from preclinical models translate to humans. Namely, the repetition suppression (but not deviance detection) in humans is GABA-dependent, and is disrupted in ASD. Our results go beyond and suggest that arbaclofen ameliorates repetition suppression anomalies in autistic adults.

### Heterogeneous neurophysiological responses to arbaclofen – clinical relevance

Activating GABA receptors through arbaclofen has been suggested as a potential therapeutic strategy for ASD (*55*) and fragile X syndrome (FXS) (*66*), among the most common single-gene causes of ASD. Clinical Trials of arbaclofen did show some promise in subgroups, but failed to change the primary outcome measure overall (*55*). Our results also show heterogeneity in the neurophysiological response to GABA_B_ agonism in ASD. The reasons for heterogeneous responses to arbaclofen are likely to be complex. The metabotropic GABA_B_ receptors are conventionally thought to provide tonic inhibition and regulate cellular excitability through both pre- and postsynaptic mechanisms (*67, 68*). They also have crosstalk with the glutamate system, as well as GABA_A_ receptors. Furthermore, they may impact overall GABA production and breakdown, and likely exert broad downstream cellular effects including both inhibition and disinhibition (*69*). Thus, boosting GABAergic function through arbaclofen may affect a range of mechanisms that differentially modulate the excitatory and inhibitory targets in individuals with and without ASD.

### Cellular and developmental bases of repetition suppression differences

Though our study cannot establish exactly what cellular differences in ASD explain the weak repetition suppression there are hints from preclinical studies. The oddball paradigm has been widely used in translational studies across species to study repetition suppression or SSA and provides a number of converging observations. First, it has been demonstrated that GABAergic somatostatin-positive interneurons (SOMs) selectively suppress responses to repetitive standards but not deviants in the primary auditory cortex in mice (*11*). Second, others have found that silencing SOMs in rodents leads to loss of rodent visual MMN (*12*). Third, SOMs show late input/output facilitation in the MMN timescale (*70*) and have a slow firing rate which is well suited to serve as substrates of the slow theta, alpha and beta rhythms supporting neuro-oscillatory responses to repeated stimuli (*49*) and the activity of SOMs is associated with cortical oscillations within these frequency bands (*71, 72*). Thus, a candidate mechanism underpinning the observed weaker repetition suppression in ASD and the differential GABAergic modulations by arbaclofen in TD and ASD may be altered interaction of SOMs with the pyramidal neurons in local circuits along the hierarchical auditory pathways.

Preclinical examination of SSA suggests that postsynaptic GABA_B_ receptor activity is needed to reduce pyramidal response to repeated sounds, whereas presynaptic GABA_B_ receptor activity promotes responses to repeated sounds (*40*). Thus, speculatively, the postsynaptic GABA_B_ receptors response is dominant in suppression in TD. Excessive GABA_B_ receptors activation with arbaclofen may shift the typical balance towards presynaptic GABA_B_ receptor activity and disrupt suppression. In ASD, since arbaclofen increases repetition suppression, this could indicate that postsynaptic GABA_B_ receptor mechanisms are altered at baseline; but further experimental work in animal models will be needed to test this concept.

### Origins of GABA differences in ASD

Although we carried out our study in adults, ASD is a neurodevelopmental condition that has its origins in the early life. Sensory circuits in which GABA has a key role mature in early postnatal periods and subsequent brain development ‘cascades’ through multiple sensitive periods as more and more complex cognitive and behavioral skills are acquired (*73*). Indeed, atypical sensory processing has been flagged very early in development of infants who go on to receive a diagnosis of ASD (*2, 74*). Thus, the atypical GABAergic auditory processing in ASD observed here is likely to reflect earlier alterations in neural circuit maturation. Consistent with this, others have reported altered auditory cortical reactivity in newborn infants at high-risk of ASD (*2, 57*). These GABAergic developmental perturbations are unlikely to be restricted to the auditory domain. We have documented tight links between the GABA system and altered tactile processing in ASD (*15*) and GABA-dependent differences in fundamental visual processing in ASD which are also restored to a more typical pattern with arbaclofen (*13*). Prospective longitudinal studies will help map causal pathways from sensory abnormalities to later emerging symptoms in ASD.

### Sensory processing and wider autistic symptoms

It was also beyond the scope of our study to examine how GABAergic modulation alters sensory *symptoms* (as opposed to sensory processing) in ASD, however, the strong correlation between the individual sensitivity index and AQ scores observed here supports the concept that the neuropathology underpinning atypical sensory suppression link to a wider phenotype.

Individuals with ASD often report that overwhelming sensory stimulation makes it difficult to interact with the environment (*3*). Differences in sensory processing have been identified across neurophysiology, perception and emotional responses to sensory inputs. Overwhelming sensory experiences have also been suggested to contribute to the anxiety which is extremely common in ASD, and predispose individuals with ASD to repetitive behaviors (*75*). Thus, sensory processing differences have tremendous impact in themselves and have knock-on effects on mental health and behavior. Our results suggest that the neurophysiology of auditory processing is linked with both sensitivity to sounds and wider phenotypes across our cohort as captured using the AQ. We show that the extent to which GABA-dependent neuro-oscillatory response (ERSP) to repeated auditory stimuli is atypical in ASD and related to an individual’s phenotype. This is potentially important because the neuro-oscillatory activity provides an essential platform for functional connectivity across brain networks (*76*). Therefore, our results raise the possibility that altered GABA-regulation of neuro-oscillatory responses to sensory stimuli has ‘knock-on’ effects across brain networks and the complex cognitive and behavioral functions they subserve. Investigations of such ‘fundamental’ sensory processes in ASD may therefore provide a more accessible route to modulating downstream difficulties experienced by autistic people.

We emphasize that our findings do not speak to the clinical efficacy of arbaclofen but may have implications for the development of pharmacological interventions targeting core ASD symptomatology. In clinical trials of candidate drugs developed for ASD, primary endpoints generally rely upon various behavioral checklists, as for example the Autism Diagnostic Observation Schedule, the Social Responsiveness Scale, the Autism Diagnostic Interview-Revised and other similar tests. Such behaviors are highly complex: they are generally established on multiple lower-order processes and shaped by variable gene-environment interactions throughout life. The results rely on subjective responses from either participants or their care providers. In contrast, our findings indicate that dynamic changes in sensory processing can be passively measured and quickly respond to a single dose of candidate drugs. A next step would be to explore whether individual sensory responses are useful pretrial to identify whether individuals who are biologically responsive to a candidate drug a candidate drug are also clinically responsive. This may help promote more personalized, mechanism-informed and cost-efficient approaches to clinical trials.

### Arbaclofen side effects

Known side effects of arbaclofen such as dizziness or nausea were reported by some participants occasionally throughout the study, but as expected, more so in the high-dose condition. For example, three participants with ASD reported fatigue and one TD participant reported for dizziness which were classified as moderate because they were more than ‘mild’. Mild side effects were very minimal, essentially any passing mention of side effects reflected a clear comment from the participant of a noticeable experience. This was not evaluated in terms of impact on function. These observed side effects could still potentially have limited acute dose studies by affecting the participants’ attentional resource allocation during the task. However, the auditory oddball paradigm used in this study was a passive paradigm with minimal demands on attention. The predictive response persists irrespective of attention during sleep (*77*) and anesthesia (*78*) and is detectable in early infancy (*79*), making it a promising tool for investigations of neurodevelopmental disorders (*80, 81*). We hope that this approach has a potential for generalization to individuals across age groups with immature or impaired cognitive abilities that are frequently excluded from drug development studies.

### Conclusions

In this study of GABAergic modulation of auditory processing we demonstrated weaker repetition suppression in ASD compared to TD. We show, for the first time in humans, that activating GABA_B_ receptors through a single oral dose of arbaclofen shifts atypical auditory processing in ASD towards a more typical profile; and disrupts typical responses in TD. Thus, we have directly confirmed that GABAergic dysfunction contributes to the neurophysiology of auditory sensory alterations in ASD.

## Materials and Methods

### Study design

Participants provided written informed consent for an experimental medicine study comprising a series of experiments to investigate the role of GABA on ASD (*13*) approved by King’s College Research Ethics Committee (RESCM-17/18-4081). Participants were given placebo or a single oral dose of 15 or 30 mg of arbaclofen (STX209) on the study day, three hours before the auditory task. The order of administration of study drug or placebo was randomized to prevent order effects. Arbaclofen plasma concentration is expected to have a half-life of five hours (*82*); thus, the testing was within the active physiological window. All the tests were conducted at the Institute of Psychiatry, Psychology and Neuroscience, King’s College London. Study visits were at least one week apart to ensure complete drug wash-out.

Medical cover was provided throughout each test session, and participants were asked to remain at our unit at least four hours after drug/placebo intake. The medic was ‘blind’ to the order of administration but had access to the allocation information held at our pharmacy and by the Chief Investigator if needed. No emergency access to allocation was required, but where a participant had experienced side-effects which were more than moderate in the opinion of the study clinician and after discussion with the Chief Investigator, unblinding occurred to try to avoid exposure to a higher dose of arbaclofen on a subsequent visit.

### Participants

All participants were adults, aged from 19 to 53, with intelligence quotient (IQ; on the Wechsler Abbreviated Scale of Intelligence; WASI-II (*83*)) > 70. Demographic characteristics including biological sex and IQ did not differ between the TD and ASD groups (Table 1). The mean age of the ASD group (34.8 years) was higher than that of the TD group (28.6 years). We added age as a fixed variable in the LMM model and re-ran the statistical analysis. The results showed there was no effect of age on any temporal or spectral responses measured; thus, age difference was unlikely to explain any group differences reported. For the ASD group, 16 participants were recruited from our National Autism and ADHD service for Adults (NAASA) at the South London and Maudsley National Health Service (NHS) Foundation Trust; the service protocol includes an Autism Diagnostic Interview-Revised (*84*) where an appropriate informant is available and/or Autism Diagnostic Observation Schedule (*85*) to inform the current symptom level (www.slam.nhs.uk/national-services/adults-services/adhd-and-asd-services-outpatients/). However, for inclusion in the ASD group of this study, we required a clinical diagnosis to be in place i.e. the final diagnostic decision was a clinical opinion made by an experienced psychiatrist or clinical psychologist from in a recognized ASD multidisciplinary clinical assessment setting in the UK. Those individuals recruited outside of our National Autism and ADHD Service for Adults (NAASA) at the South London and Maudsley NHS Foundation Trust were carefully screened by an experienced NAASA clinician for inclusion. The clinician had to be satisfied by the account of their diagnostic assessment through a recognized U.K. autism service (with documentary evidence where possible), in addition to their responses to our screening interview. ASD traits were also assessed across both the TD and ASD group using the AQ. The results showed a highly significant group difference in AQ as expected (t = 9.1, p = 7×10^−13^; Table 1). We also extracted responses to AQ question 5: “I often notice small sounds when others do not” and coded “strongly disagree; somewhat disagree; somewhat agree; strongly disagree” from 1 to 4 respectively.

Participants with ASD with a known genetic cause, such as fragile X syndrome, neurofibromatosis type 1, or 22q11 deletion syndrome, were excluded from the study. Other inclusion criteria were as follows: ability to give informed consent, no comorbid psychiatric illness such as psychotic illness and major mood disorder, no history of seizures or diagnosis of epilepsy, and no physical illness, such as heart disease, high blood pressure, and renal insufficiency. In the month preceding participation, 13 participants (3 TD and 10 ASD) were taking regular medication with drugs such as sertraline, ibuprofen and citalopram, which did not affect glutamate or GABA directly. All other participants were medication-free.

### Auditory oddball paradigm

On each study visit, a total of 1,400 auditory stimulus trials were presented to participants in a classical oddball paradigm (*53*). The stimulus train comprised a sequence of four different types of sinusoidal tones: 82% standard trials (1000 Hz, 50 ms), 6% frequency deviants (1200 Hz, 50 ms), 6% duration deviants (1000 Hz, 100 ms) and 6% combined frequency-duration deviants (1200 Hz, 100 ms). The sequence order was initially generated by a random function in MATLAB 9.2.0 (MathWorks Inc.) and remained consistent across participants. The inter-stimulus interval (ISI) was a randomized value between 500 and 600 ms. The sounds were presented at 70 decibels (dB) via speakers in an enclosed room. Participants were comfortably seated and instructed to watch a muted movie to distract their attention from the sounds during the test.

### EEG acquisition and pre-processing

Continuous scalp EEG signals were recorded via a 64-channel standard actiCAP (EASYCAP GmbH) with a sampling rate of 5 KHz and amplified by a BrainAmp amplifier (Brain Products GmbH). The default reference was FCz and electrode placements followed the international 10-20 system. Impedances between the scalp and electrodes were kept below 15 kΩ.

Offline pre-processing was conducted using custom scripts that included functions from the EEGLAB (*86*) in MATLAB. Raw EEG data was first re-referenced to the mean of mastoids, down-sampled to 250 Hz and filtered between 0.1 and 30 Hz using zero-phase finite impulse response filters. Next, continuous EEG was data segmented for each stimulus trial in the interval [-100, 500] ms referenced to the stimulus onset at 0 ms. The Hyvärinen’s fixed-point algorithm (*87*) for independent component analysis (ICA) was performed by calling the ‘fastICA’ function to visually detect potential artifacts. A single channel (FC1) over the front-central region was selected for further analysis because MMN was reported to reach maximum amplitude in this area (*20*). Trial epochs with voltages exceeded ± 100 μV were regarded as contaminated by eye blinks or unexpected artifacts and excluded from analysis.

### ERP analyses of MMN

As the number of standard trials was far exceeded the number of deviant trials in the oddball paradigm, we randomly selected a subset of standard trials to reach a balanced number of standard and deviant stimulus conditions in each study visit. The size of the standard subset equaled the mean of acceptable deviant trials. Next, the pre-processed trial epochs were averaged as a function of stimulus condition. Three difference waves (frequency, duration and frequency-duration) were calculated by subtracting the average standard response from the average response to the corresponding deviant, respectively. Further, difference waves were separately averaged for the TD group and the ASD group at each drug condition to achieve the grand average ERP waveforms, as shown in Fig. 1A. MMN was defined to be the negative peak that appeared usually within [100, 200] ms post stimulus onset (*88, 89*). In this study, at placebo, the grand average latencies of the frequency MMN and the frequency-duration MMN were in the [100, 200] ms window as expected, whilst the duration MMN was prolonged to locate at [200, 300] ms (Fig. 1A). To improve consistency in the analysis, we defined a time window [100, 300] ms regardless of the stimulus condition to measure individual MMN ERP features including the peak amplitude and its corresponding latency. Please see Fig. 1B for the individual-level scatter plots of MMN ERP features.

### ERP analyses of responses to standard and deviant tones

In addition to the difference waves, we also examined neural responses to each stimulus condition without subtraction. The grand average ERP waveforms as a function of the stimulus condition and the drug administration for TD and ASD are shown in Fig. 2A. We focused on two ERP components that were prominent in the grand average waveforms: (i) The P1 (P50) component appeared as a positive peak within [50, 100] ms post stimulus onset; (ii) The N1 component followed P1 as a negative valley in the range [100, 200] ms. In response to the duration deviant, there was another negative valley (N2) located in the range of [200, 300] ms, which maintained an amplitude similar to the N1 component evoked by the frequency deviant and the combined deviant, larger than the N1 component of the duration deviant. Moreover, the temporal range of this N2 component was more aligned with that of the duration MMN as shown in Fig. 2A. Thus, the window for individual measurement of the negative component of the duration deviant was set as [200, 300] ms (instead of [100, 200] ms for other stimuli) to ensure we were measuring the same neural component that contributed to the MMN generation across stimulus conditions. The amplitudes of the P1 and N1 were then extracted for each participant for statistical analysis as shown in Fig. 2B.

### Time-frequency analyses of responses to standard and deviant tones

We used ERSP analysis to measure dynamical changes in spectral bands after stimulus onset. The balanced trial epochs used in the ERP analyses were fed as input to the ERSP function ‘newtimef’ implemented with EEGLAB. Specifically, the time limits of the trial epochs were [-100, 500] ms referenced to the stimulus onset with the sampling rate of 250 Hz. The frequency limits were set as [4, 25] Hz with 0.5 Hz frequency resolution. Baseline spectra were calculated from the pre-stimulus interval [-50, 0] ms. Each epoch was then divided into overlapping Hanning windows with a range of 128 ms (32 samples) and a step-size of 4 ms. Spectra of each window was normalized with reference to the baseline and assigned to the center point of the window. Time limits of the output ERSP were [-36, 436] ms referenced to the stimulus onset. Normalized transforms of repeated trial epochs were then averaged by stimulus conditions and recorded as log values in a time-by-frequency ERSP matrix.

Grand average ERSP outcomes of participants grouped by the drug administrations were calculated by averaging ERSP matrixes in each group. As shown in Fig. 3A, the main event-related perturbations at the grand average level occurred within the theta and the alpha band [4, 12] Hz. Therefore, for each participant visit, ERSP values in the [4, 12] Hz band were summed up to obtain the evoked power waveform. The mean of the evoked power waveform within specified time window was extracted as a single-point measurement of the event-related spectral dynamics. The measurement windows used for the four stimulus conditions were the same as the N1 settings in the ERP analyses ([200, 300] ms for the duration deviant, [100, 200] ms for others). Please see Fig. 3B for the scatter plots of individual ERSP results.

### Comparisons between responses to pre-deviant standards and post-deviant standards

We applied both ERP and ERSP analyses to compare data epochs obtained from the pre-deviant standard trials and those from the post-deviant standard trials. The pre-deviant standard was defined to be the last sound in a four-in-a-row standard sequence before a deviant; the post-deviant standard was the first standard after a deviant with an interval of one second. The numbers of accepted trials were balanced between the pre- and post-deviant standards (96 trials per participant for each type). The results of ERP and ERSP analyses were shown in Fig. 4. Individual ERSP was measured as the mean of a 50-ms interval around the average peak perturbation within the range of interest [50, 150] ms post stimulus onset. Other parameters used for the ERP and ERSP analyses were the same as the settings applied on standard responses in previous sections.

To measure how a single individual responded to arbaclofen, we defined a sensitivity index as the placebo-30 mg difference in spectral responses to pre-deviant standards. We calculated the index for the 29 participants (12 TD, 17 ASD) that completed both the placebo and 30 mg visits and ran an independent-sample t-test to confirm the group difference. Further, a correlation analysis between the sensitivity index and quantified ASD traits measured by the AQ (total score and score of response to question 5) was done to investigate the relationship with general ASD phenotype and sound perception, respectively.

### Statistical analysis

For ERP and ERSP individual outcomes to different stimuli, we first used independent-sample *t* test to assess ASD-TD group differences in responses to each stimulus condition at placebo and drug administrations. The p-values were corrected for multiple comparisons using the Benjamini-Hochberg method.

The linear mixed-effect model (LMM) was applied with the stimulus, drug dose and group as fixed-effects variables. The stimulus condition was categorical and others were numeric. The LMM ran pair-wise comparisons within categorical stimuli, providing direct measurements of repetition suppression between responses to the standard tone and deviants. Next, pure numeric LMM models were built for analyses of responses to a specific stimulus such as the standard tone, in which drug dose and group were set as fixed-effects variables. The latter was regarded as more interpretable because it provided a measure of differential modulations by arbaclofen in TD and ASD reflected as the group-drug interaction. When a significant interaction was observed, the LMM model further shrank to be a simple linear model with drug dose as the independent variable and was separately applied to TD and ASD to measure the drug effect. At each stage, the p-values were corrected for multiple comparisons using the Benjamini-Hochberg method.

## Data Availability

Data from this study are available on request.

## Acknowledgements

The authors thank all the participants for their support. They also thank Paul Wang, Luke Mason, Johanna Kangas, Cornelia Carey, Charlotte Pretzsch, Alison Leonard and Beata Kowalewska for their help with study logistics. This project was funded by an Independent Investigator Award (G.M.M.) from the Brain and Behaviour Research Foundation and by Clinical Research Associates, L.L.C. (CRA), an affiliate of the Simons Foundation. Support is also acknowledged from Autistica (A.C.P.) and the Sackler Institute for Translational Neurodevelopment at King’s College London and EU-AIMS (European Autism Interventions)/EU AIMS-2-TRIALS, an Innovative Medicines Initiative Joint Undertaking under Grant Agreement No. 777394. In addition, this paper represents independent research part funded by the National Institute for Health Research (NIHR) Mental Health Biomedical Research Centre (BRC) at South London and Maudsley NHS Foundation Trust and King’s College London. The views expressed are those of the author(s) and not necessarily those of the NHS, the NIHR or the Department of Health and Social Care.

## Author contributions

G.M.M. conceived the study. Q.H., H.V., A.C.P., J.A., E.D., G.M.M. and D.G.M.M. designed the study. Q.H., G.M.M., S.F.C, N.A.J.P., D.B., H.V., A.C.P., J.A. and G.I. advised on the methods. H.V., A.C.P., C.L.E, F.M.P., N.M.L.W., E.D., L.K. and M.D. collected the data. Q.H., H.V. and J.A. preprocessed the data. Q.H. analyzed the data. Q.H. and G.M.M. drafted the manuscript. All authors edited and approved the final draft.

## Data availability

Data from this study are available on request.

